# Integration of urine proteomics and renal single-cell genomics identifies an interferon-gamma response gradient in lupus nephritis

**DOI:** 10.1101/2020.03.13.20034348

**Authors:** Andrea Fava, Jill Buyon, Chandra Mohan, Ting Zhang, H. Michael Belmont, Peter Izmirly, Robert Clancy, Jose Monroy Trujillo, Derek Fine, Yuji Zhang, Laurence Magder, Deepak A. Rao, Arnon Arazi, Celine C. Berthier, Anne Davidson, Betty Diamond, Nir Hacohen, David Wofsy, the Accelerating Medicines Partnership in SLE network, Soumya Raychaudhuri, Michelle Petri

**Affiliations:** Division of Rheumatology, Johns Hopkins University School of Medicine, Baltimore, MD, USA; New York University School of Medicine, New York, New York, USA; University of Houston, Houston, USA; Division of Nephrology, Johns Hopkins University, Baltimore, MD, USA; Department of Epidemiology and Public Health, University of Maryland, Baltimore, MD, USA; University of Maryland Marlene and Stewart Greenebaum Comprehensive Cancer Center, Baltimore, Maryland; Division of Rheumatology, Immunology, Allergy, Department of Medicine, Brigham and Women’s Hospital, Harvard Medical School, Boston, MA, USA; Broad Institute of MIT and Harvard, Cambridge, MA, USA; Internal Medicine, Department of Nephrology, University of Michigan, Ann Arbor, MI, USA; Center for Autoimmune and Musculoskeletal Diseases, The Feinstein Institute for Medical Research, Northwell Health, Manhasset, NY, USA; Rheumatology Division, University of California San Francisco, San Francisco, CA, USA; Multiple Organizations; Center for Data Sciences, Brigham and Women’s Hospital, Boston, MA 02115, USA; Division of Rheumatology and Genetics, Department of Medicine, Brigham and Women’s Hospital, Boston, MA 02115, USA; Department of Biomedical Informatics, Harvard Medical School, Boston, MA 02115 USA; Program in Medical and Population Genetics, Broad Institute of MIT and Harvard, Cambridge, MA, USA; Centre for Genetics and Genomics Versus Arthritis, Centre for Musculoskeletal Research, Manchester Academic Health Science Centre, The University of Manchester, Oxford Road, Manchester, UK

**Keywords:** systemic lupus erythematosus, lupus nephritis, proteomics, single cell transcriptomics, interferon-gamma

## Abstract

Lupus nephritis, one of the most serious manifestations of systemic lupus erythematosus (SLE), has both a heterogeneous clinical and pathological presentation. For example, proliferative nephritis identifies a more aggressive disease class that requires immunosuppression. However, the current classification system relies on the static appearance of histopathological morphology which does not capture differences in the inflammatory response. Therefore, a biomarker grounded in the disease biology is needed to understand the molecular heterogeneity of lupus nephritis and identify immunologic mechanism and pathways. Here, we analyzed the patterns of 1000 urine protein biomarkers in 30 patients with active lupus nephritis. We found that patients stratify over a chemokine gradient inducible by interferon-gamma. Higher values identified patients with proliferative lupus nephritis. After integrating the urine proteomics with the single-cell transcriptomics of kidney biopsies, it was observed that the urinary chemokines defining the gradient were predominantly produced by infiltrating CD8 T cells, along with natural killer and myeloid cells. The urine chemokine gradient significantly correlated with the number of kidney-infiltrating CD8 cells. These findings suggest that urine proteomics can capture the complex biology of the kidney in lupus nephritis. Patient-specific pathways may be noninvasively tracked in the urine in real time, enabling diagnosis and personalized treatment.

## INTRODUCTION

Lupus nephritis is one of the most severe manifestations of systemic lupus erythematosus (SLE) (1). Lupus nephritis affects up to 50% of patients leading to end-stage renal disease in 10% and carries an 8-fold increase in mortality (2–8). Since nephritis is usually asymptomatic, patients with SLE are serially screened for the presence of abnormal urine protein (9). When proteinuria is elevated, a renal biopsy is obtained to confirm the diagnosis and guide treatment. Histopathology classifications subgroup lupus nephritis into six classes based on the presence, amount, and location of inflammation and fibrosis (10). For example, the presence of intraglomerular immune cell infiltration (class III and IV) identifies the most aggressive proliferative form of SLE nephritis, which is treated with stronger immunosuppression. Membranous glomerulonephritis (class V) is characterized by thickened glomerular basal membranes, subepithelial immune complex deposition and absence of intraglomerular infiltration. Endocapillary proliferation and membranous disease frequently overlap in the “mixed” phenotype.

Although the current treatment approach to lupus nephritis is loosely grounded in the morphological classification, findings from recent studies have challenged this paradigm. Kidney biopsies repeated after one year showed a transition in class in up to 70% of cases, from non-proliferative to proliferative and vice versa, with the second biopsy having better prognostic value (11, 12). These findings indicate that lupus nephritis is a dynamic process and therefore disease state inferred from kidney biopsies at one time point may have a limited value. In addition, recent studies have underscored the prognostic importance of interstitial disease which is inadequately addressed by current histologic classification schemes (13). Finally, routine histopathology of lupus nephritis cannot evaluate the underlying molecular pathways that may inform personalized treatment choices. Recent single-cell RNA-seq studies have revealed the complex network of immune cells, and their diversity, in the context of lupus nephritis (14–16). These studies offer promise for more personalized strategies for therapeutics and for prevention of renal fibrosis.

As patients with the same histological class may have dramatically different outcomes (17), there is an unmet need for a new biology-based classification of lupus nephritis that can dissect the heterogeneity. Ideally, such immunologic classification needs to be easily queried during the course of the disease to dynamically assess changes in response to intervention over months (rather than year). Several previous studies explored potential biomarkers to assess and predict lupus nephritis by using targeted and systematic proteomic approaches (18–21). However, these have not yet been standardized for clinical practice and do not consider lupus nephritis biological endotypes (18). In this study, we sought to identify patterns in the urine proteome that can identify distinct groups of SLE patients and infer the ongoing renal molecular pathology. We quantified 1000 urine protein biomarkers in active lupus nephritis and identified that patients stratify on an immune activation gradient. Integration with renal single-cell RNA sequencing revealed that the urine signature reflected intrarenal secretion by immune infiltrating cells and that interferon-gamma is a main driver of such increased immune activation.

## RESULTS

### Urine proteomics stratifies lupus nephritis patients on an immune activation gradient

This study was initiated to address whether we could classify lupus nephritis patients based on distinct biological patterns in the urine. We quantified 1000 analytes including cytokines, growth factors, and other soluble markers in the urine from 30 SLE patients with proteinuria and a kidney biopsy on the same day confirming lupus nephritis (class III, IV, or V) as part of Phase 1 of the Accelerating Medicine Partnership (AMP; **Table 1 and figure S1**) (14). To agnostically determine patterns of inflammation, we used principal component analysis (PCA), a dimension reduction technique that identifies major axes of variation in the urine proteome. For example, the first two principal components (PCs) clearly separated healthy donors from SLE patients based on distinct patterns of urinary protein excretion (**Figure S2**). Within lupus nephritis, the first principal component (PC1) stratified patients on a gradient rather than distinct clusters (**Figure 1A**). We noted that patients with higher PC1 values were more likely to have proliferative lupus nephritis, while the patients with pure membranous lupus nephritis had PC1 values close to 0 or negative (**Figure 1A-B**) suggesting that PC1 detected a biological response shared by many patients with proliferative lupus nephritis. No association was detected between PC1 and demographics or technical confounders such as batch, proteinuria, age or gender. PC2 was associated with site of urine collection (**Figure S3)** and therefore it was not evaluated further.

**Table 1.** Demographics and clinical characteristics.

| Characteristic | n (%) / mean [range] |
| --- | --- |
| Age | 33.5 [19-54] yrs |
| Female | 28 (93%) |
| Race/Ethnicity |  |
| African American | 12 (40%) |
| Caucasian | 10 (33%) |
| Asian | 7 (23%) |
| Other | 1 (3%) |
| Anti-dsDNA | 24 (80%) |
| Low complement | 26 (87%) |
| Lupus nephritis class |  |
| III or IV | 14 (47%) |
| V | 9 (30%) |
| III+V or IV+V | 7 (23%) |
| Histological features (n=19) |  |
| Activity | 3 [0-12] |
| Chronicity | 1 [0-4] |
| Proteinuria, mg/g | 2.6 [0.53-11.6] |
| Response to treatment at 12 months | 6 (20%) |
| Treatment at time of biopsy |  |
| Hydroxychloroquine | 26 (87%) |
| Corticosteroids | 19 (63%) |
| Mycophenolate | 1 (3%) |
| Belimumab | 2 (7%) |

**Figure 1.**
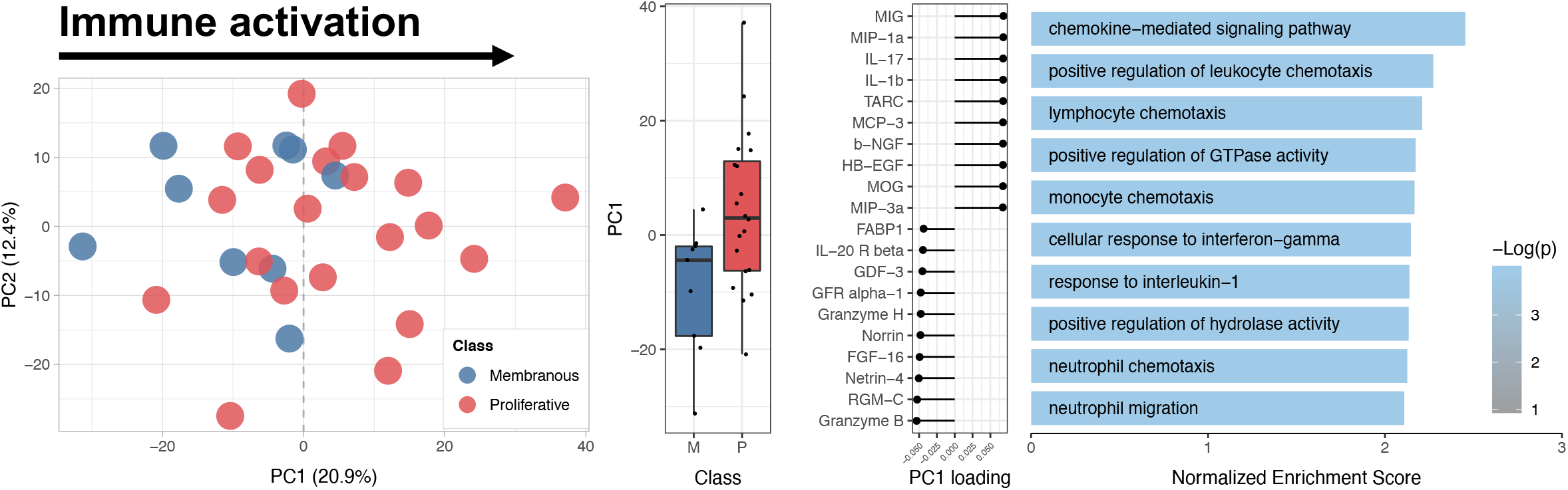
Lupus nephritis patients stratify over an immune activation gradient. (A-B) PCA (n=30) of the first 2 principal components of the urine proteome (% variance explained is indicated). Patients with higher PC1 value have almost exclusively proliferative lupus nephritis (class III, IV, or mixed). P value calculated by t test. (C) Top and bottom 10 PC1 loading values of the measured urine protein. (D) Top 10 enriched pathways PC1 using Gene Ontology Biological Process indicating the biological significance of PC1.

Next, we sought to characterize the biological significance of the PC1 gradient. As principal components are determined by the weighted sum of all the analytes, we tested whether a molecular pathway was enriched using the weight (“loading”) of each analyte on PC1 (**Figure 1C**). Gene set enrichment analysis (GSEA) revealed that PC1 detected chemotaxis pathways (FDR < 0.01) (**Figure 1D and S4**): in particular, a pattern of chemokines secreted in response to IFN-gamma, IL1-beta, and TNF and directed to attract monocytes, NK, and CD8 T cells.

Together, these findings indicate that urine proteomics analyses identified a differential expression of a specific immune activation signature in patients with lupus nephritis and that stronger signals were associated with proliferative lupus nephritis.

### Urine cytokines reflect intrarenal production by myeloid and cytotoxic lymphocytes

We asked whether the chemokines detected in the urine were indicative of intrarenal production by kidney infiltrating immune cells. To answer this question, we analyzed single-cell transcriptomics from 24 renal biopsies of patients with lupus nephritis (**Figure 2A-B**) (14). We carried out single-cell transcriptomic analysis on genes coding for the proteins found in two of the most enriched pathways in the urine PC1 (Chemokine-mediated signaling pathway GO:0070098 and Cellular response to interferon-gamma (GO:0071346)) (**Figure S5**) in the renal cells. A chemokine score was defined as the sum of the normalized expression of the aforementioned genes. While these chemokines were expressed by most infiltrating immune cells as well as epithelial cells, the dominant expression was observed in myeloid, NK, and CD8 T cells (**Figure 2C-F**).

**Figure 2.**
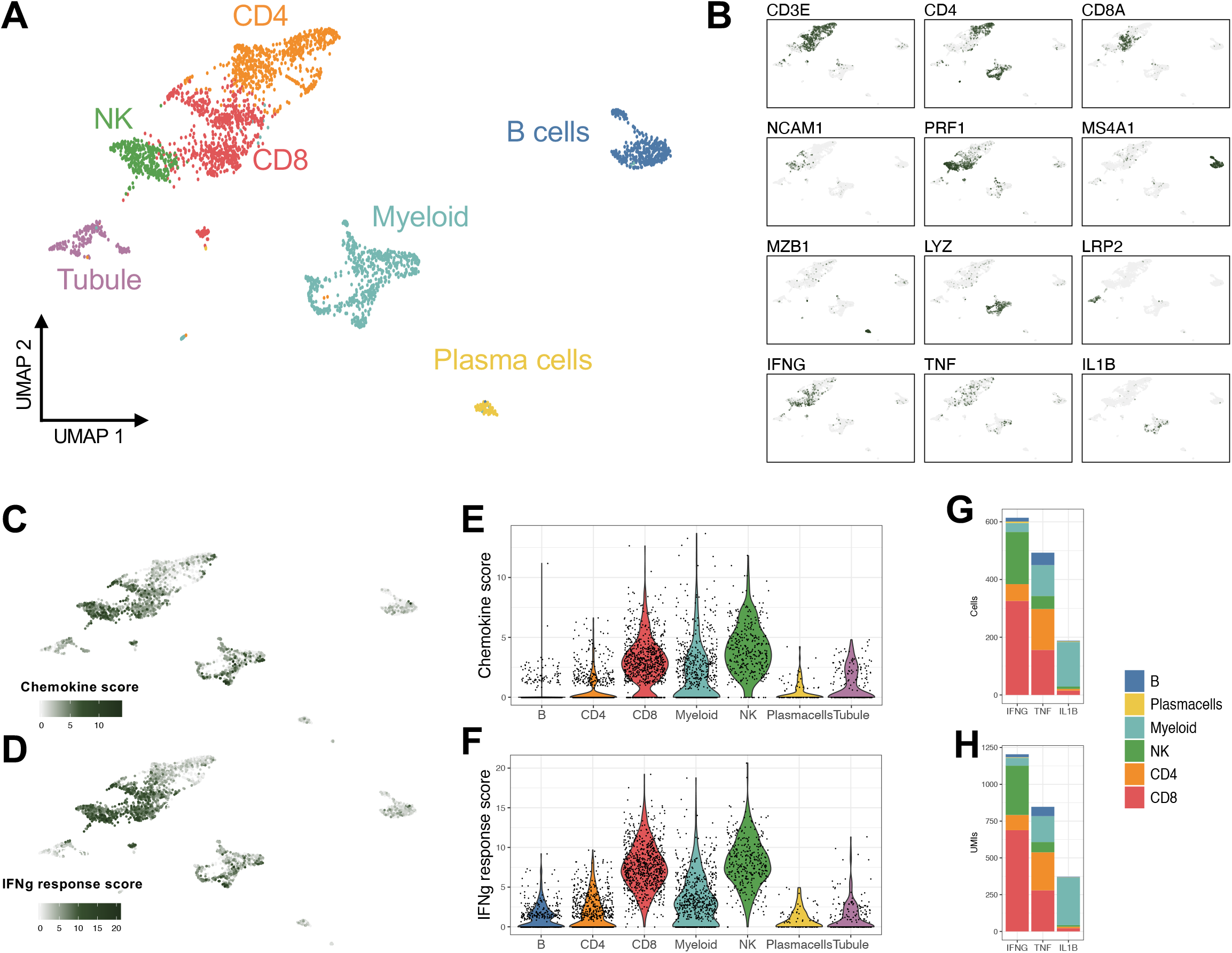
The chemokine pathway detected by urine proteomics reflect intrarenal production by myeloid, CD8, and NK cells. (A) UMAP plot of single-cell RNA-sequencing of 24 renal biopsies of patients with active lupus nephritis (medium resolution clustering). (B) The identity of the cell clusters was determined by the expression of lineage markers. (C-F) A chemokine and an IFNγ response score based on the expression of the markers in the top urine PC1 pathways (“Chemokine-mediated signaling pathway GO:0070098” and “cellular response to interferon-gamma (GO:0071346)”) identified kidney infiltrating myeloid, CD8, and NK cells as the main producers of the chemokines in response to IFNγ detected in the urine. (G) Distribution of IFNγ, TNFα, and IL1β expressing cells. (H) Distribution of the total UMIs mapped to IFNγ, TNFα, and IL1β as a proxy for the amount of cytokine produced by each cell type.

To evaluate whether the proteins found in the urine derived from serum as a consequence of glomerular or tubular damage, we repeated the analysis after adjustments for albumin or beta 2 microglobulin, respectively. Results were virtually identical to the unadjusted analysis (**Figure S6**).

These findings suggest that the chemokines identified by urine proteomics derive from intrarenal chemokine production and, in particular, by myeloid, NK, and CD8 T cells.

### IFN-gamma is mostly produced by infiltrating CD8 and NK cells in all patients with lupus nephritis

Pathway analysis indicated a chemokine pattern inducible by IFN-gamma, IL1-beta, or TNF. We quantified the kidney-infiltrating cells expressing such cytokines. Out of the three cytokines, IFN-gamma-positive cells were the most abundant, followed by TNF and IL1-beta (**Figure 2G**). IFN-gamma was mostly produced by CD8 T and NK cells, TNF by myeloid, CD8 and CD4 T cells, and IL1-beta almost exclusively by myeloid cells (**Figure 2B, 2F, and 2I**). Local transcription of these cytokines suggests that their transcriptional signatures in the kidney and protein levels in the urine are a direct consequence of intrarenal production. Since patients contributed a different number of cells to the dataset, we evaluated whether these findings were biased by a handful of patients contributing more cells. IFN-gamma+ cells were identified in all patients ranging between 4% and 45% of the total immune cells (mean 18%), TNF+ cells in 4-32% (mean 16%), and IL1-beta+ in 0-13% (mean 7%) (**Table 2 and Figure S7**). In contrast, we did not detect any transcription of type 1 interferons (**Figure 3**) or type 2 immunity (Th2) cytokines such as IL4, IL5, or IL13 (**Figure S8)**, suggesting that a type 1 (Th1) immune response (22) is ubiquitous in all patients with lupus nephritis including non-proliferative histological classes and that IFN-gamma (type 2 interferon) is the main interferon being produced by immune cells in lupus kidney disease.

**Table 2.** Prevalence cytokine positive cells based on single cell RNA sequencing.

| **Class** | **IFNG** | **TNF** | **IL1B** | **Cell (n)** |
| --- | --- | --- | --- | --- |
| [III] | 17.0% | 15.0% | 0.9% | 440 |
| [III] | 12.5% | 10.1% | 4.7% | 297 |
| [III] | 10.8% | 10.8% | 3.3% | 120 |
| [III][V] | 18.2% | 15.3% | 3.5% | 170 |
| [III][V] | 16.7% | 22.2% | 0.0% | 18 |
| [IV] | 24.7% | 25.8% | 0.0% | 97 |
| [IV] | 30.0% | 32.1% | 9.1% | 243 |
| [IV][V] | 3.8% | 3.8% | 7.7% | 26 |
| [IV][V] | 13.3% | 20.0% | 6.7% | 15 |
| [IV][V] | 15.4% | 7.7% | 7.7% | 13 |
| [IV][V] | 45.0% | 16.0% | 10.4% | 338 |
| [IV][V] | 11.7% | 21.8% | 13.4% | 179 |
| [IV][V] | 22.1% | 14.0% | 2.3% | 86 |
| [IV][V] | 16.3% | 17.5% | 6.4% | 251 |
| [V] | 14.8% | 16.0% | 0.0% | 81 |
| [V] | 18.2% | 11.5% | 8.0% | 286 |
| [V] | 15.1% | 13.2% | 8.0% | 212 |
| [V] | 13.7% | 11.4% | 7.8% | 219 |

**Figure 3.**
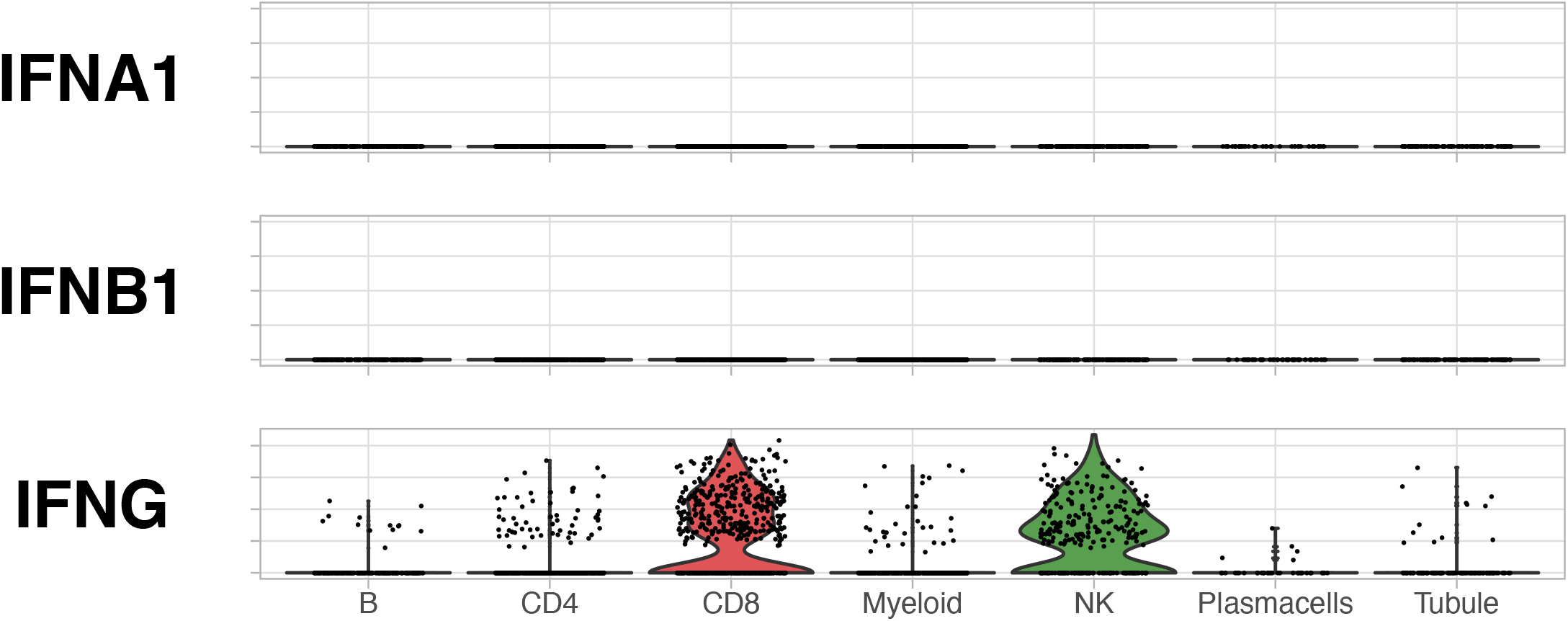
Type 2 interferon (IFN-gamma), but not type 1 (IFN-alpha or IFN-beta), is expressed in lupus nephritis kidney. Violin plots of the expression of type (IFNA1 and IFNB1) and type 2 (IFNG interferons by single cell RNA sequencing. No other type 1 interferon (i.e. IFNA2-21, IFNB3, IFNK, etc were detected (not shown).

### Urine proteomics may predict the intrarenal cellular infiltrate

We then hypothesized that urine proteomics might be used to infer the composition of kidney infiltrating cell types in lupus nephritis. We analyzed six patients with matching urine proteomics and single cell transcriptomics on a renal biopsy collected the same day. Of these, four patients were diagnosed with proliferative lupus nephritis (including mixed) and two with pure membranous lupus nephritis based on histopathology (**Figure 4A**). Single-cell transcriptomics revealed a heterogenous cellular infiltrate regardless of histopathological class (**Figure 4B**). We found that urine proteomics PC1 values strongly correlated with the relative abundance of kidney infiltrating CD8 T cells (r^2^ = 0.84, p<0.01; **Figure 4C**). There was no statistically significant correlation with the other cell populations. While needing validation in a larger cohort, these findings suggest that urinary protein patterns would predict the cell composition of the renal immune infiltrate in lupus nephritis and that CD8 T cells, the major producer of IFN-gamma, may be the key contributor of the immune activation signature in the urine.

**Figure 4.**
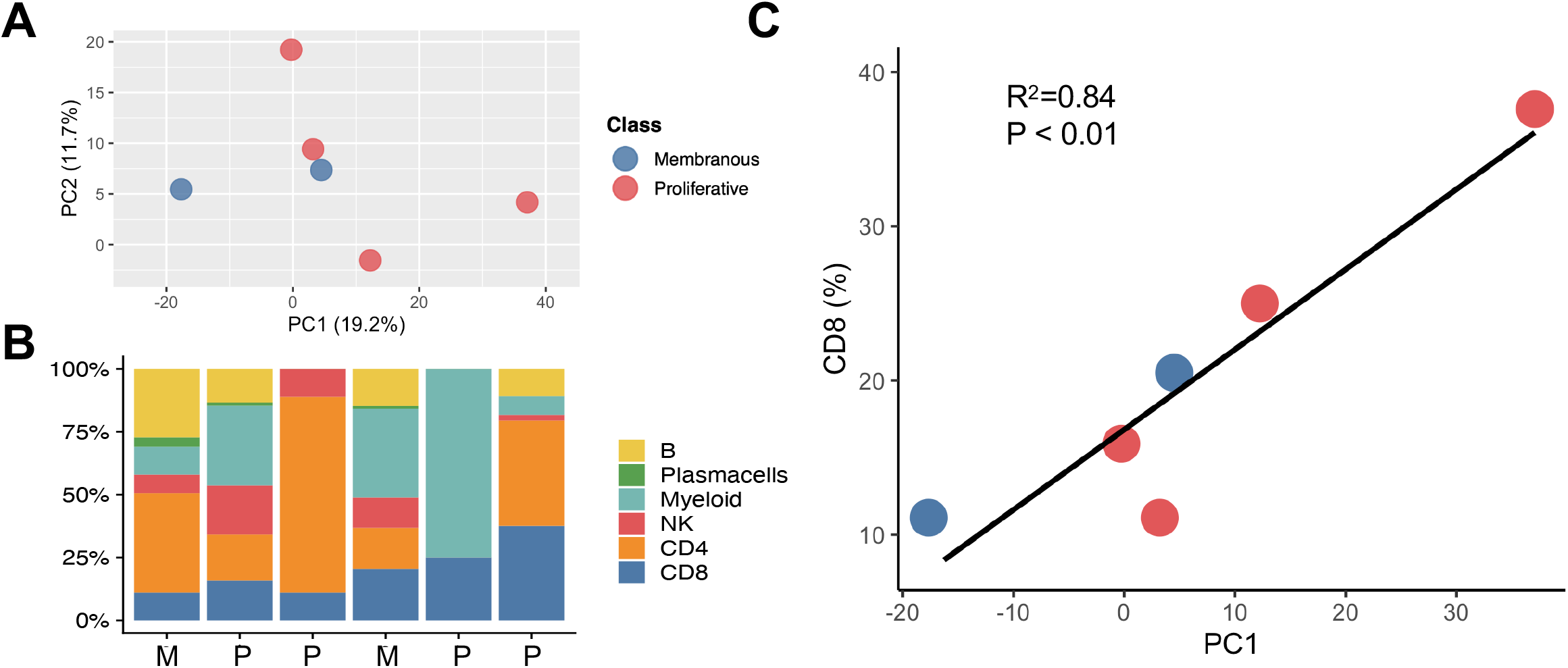
Urine proteomics may predict the intrarenal cellular infiltrate. Six patients had matching urine proteomic and single cell RNA sequencing data. Their coordinates on the urine proteomics PCA shown in Figure 1 are plotted in (A). (B) Distribution of the kidney infiltrating immune cells determined by single cell RNA sequencing. Patients are ordered based on their urine proteomics PC1 value as they appear in (A). (C) Correlation of the urine PC1 values and the frequency of kidney infiltrating CD8.

## DISCUSSION

Urine is an ideal non-invasive specimen to repetitively assess lupus nephritis. It contains the byproducts of the ongoing renal biological processes in real time, is highly non-invasive, and can be easily monitored over time. Here, we show that urine proteomics in patients with lupus nephritis may detect biologically relevant pathways that are mirrored in the kidney tissue. We found that 1) patients with higher expression of urinary chemokine and immune activation pathways have almost exclusively proliferative lupus nephritis; 2) these chemokines are renally produced by infiltrating myeloid, NK, and CD8 T cells; and 3) urine proteomics may predict the composition of the renal cellular infiltrate.

The current classification of lupus nephritis relies on morphological features that may not fully capture the complex, dynamic, and heterogeneous biology. We showed that systematic, unbiased analysis of the urine proteome may reveal the active immunological pathways in the kidney. Not surprisingly, patients with a stronger chemokine signal in the urine have glomerular immune cell infiltration as seen in proliferative lupus nephritis. Strikingly, the chemokines detected are part of a specific immune response inducible by IFN-gamma, IL1-beta, and TNF, that matches the inflammatory environment in the kidney. The significant presence of IFN-gamma positive cells suggests a strong type 1 (Th1) immune response (22). In fact, the analysis of the single-cell transcriptome of kidney infiltrating immune cells revealed IFN-gamma production in all patients, but not type 2 (Th2) response cytokines (IL4, IL5, and IL13). Furthermore, the IFN-gamma inducible urinary PC1 strongly correlated with the predominance of infiltrating CD8 T cells, the major source of IFN-gamma in lupus nephritis. Altogether, these findings suggest an ongoing IFN-gamma driven response in the kidneys of patients with lupus nephritis, which can be detected in real time in the urine by measurements of chemokines. These chemokines are directed to attract mostly macrophages, neutrophils and lymphocytes, and especially other IFN-gamma producing cells via ligation of CXCR3, CXCR4, and CCR5 that may amplify the immune response in a feedforward loop.

We deliberately performed an unsupervised analysis to detect biology-driven differences rather than looking for markers that associated with pre-specified outcomes such as histological class. While a stronger immune activation signal in the urine identified patients with proliferative lupus nephritis, a substantial group of patients classified as proliferative were grouped with patients with non-proliferative disease. Compared to those with an elevated immune activation signal, these patients may represent those whose nephritis differs in the molecular pathways involved, or those in a different stage of the inflammatory process. It is also conceivable that investigation of the relevant molecular pathways would provide a better assessment of the disease process in lupus nephritis compared to the current classification system based on glomerular endocapillary infiltration in histology. Since histological class may vary in >50% of patients on re-biopsy, the dynamic identification of precise disease driving pathways may better inform treatment selection, development of new treatments, and patient selection for clinical trials. Urine proteomics might also obviate biopsy sampling error. The next phase of AMP studies is aimed to molecularly dissect lupus nephritis and likely reclassify it based on disease biology.

Our results highlight the importance of IFN-gamma in lupus nephritis as a potential driver and coverable therapeutic target. For example, an increase in IFN-gamma activity is the first abnormality detected years before the diagnosis of SLE, preceding the appearance of autoantibodies and the dysregulation of type 1 interferons (23). IFN-gamma receptor polymorphisms are linked to the risk of developing SLE and IFN-gamma polymorphisms resulting in higher IFN-gamma production that predisposes SLE patients to develop proliferative lupus nephritis rather than membranous (24, 25). The immune response in proliferative lupus nephritis has in fact been shown to be skewed toward the Th1 axis, which is IFN-gamma predominant (24, 26, 27). In mouse models, IFN-gamma is a crucial cytokine for the development of lupus nephritis and its blockage prevents and ameliorates kidney disease as well as reduces mortality (28–30). Strikingly, type 1 interferons are not required for the generation of germinal centers and the development of lupus nephritis in some mouse models, in contrast to IFN-gamma signaling (30). IFN-gamma has a very pleiotropic effect involving cellular events implicated in lupus nephritis, including the recruitment and activation of neutrophils, CD8 and CD4 T cells, NK cells, and macrophages (31). Importantly, this molecule induces the formation of germinal centers, Tfh development and class switch in B cells (32). In the gut, IFN-gamma mediates the death of intestinal stem cells suggesting that a similar mechanism may have implications for irreversible kidney damage and fibrosis (33). In humans, encouragingly positive phase II trials showed the efficacy of blocking the IFN-gamma pathway in extrarenal lupus (34– 36). In the ustekinumab trial, for example, clinical response was associated with reduction of the IFN-gamma signature, rather than type I interferon (37). In aggregate, these findings suggest that IFN-gamma is a central cytokine in lupus nephritis and, given the acceptable safety profile of its direct blockage (38), further studies in lupus nephritis should pursue IFN-gamma inhibition.

An interferon gene response, especially type 1, is the archetypal signature of SLE (39– 41). We and others have previously showed a strong type 1 interferon signature in lupus nephritis (14–16). There is a large overlap between the gene signatures induced by type 1 and type 2 interferons (42–44), as indicated by shared signaling pathways. Each interferon type can induce the production of the other one (43) eventually leading to stimulation from both sides and thus a mixed signature. Here, we showed the unequivocal presence of IFN-gamma-producing cells in lupus nephritis along a typical chemokine signature in the urine, suggesting that IFN-gamma is central to the pathogenesis of lupus nephritis. Type 1 interferon intrarenal transcripts were not detected in our studies, suggesting that, assuming the absence of technical limitations, the source of type 1 interferons may be outside the kidney or precedes the clinical events leading to a kidney biopsy.

We acknowledge the limitations of our study. We did not detect an association with clinical variables such as renal activity/chronicity or medication use. There are likely other major detectable molecular pathways that could identify distinct patient groups. However, the relatively limited sample size did not allow for further exploration without the risk of a type 1 error. Nevertheless, our findings are confirmed by other smaller and independent studies that assessed the concentration of a fraction of our analytes in the urine of patients with lupus nephritis (45). Further studies, as those ongoing with the Phase 2 of the Accelerating Medicines Partnership, are needed to extend and validate these results. Second, while our platform did not assay the whole urine proteome, it allowed for interpretability, a broad assessment of the immune response, and limited major confounders from batch effect and unalignable peptides as commonly seen in classical mass spectrometry experiments. To limit this bias, we utilized a self-contained enrichment strategy (GSEA). In order to identify protein patterns independently of the amount of proteinuria, we have scaled the analytes concentration within and between patients. As this might introduce a bias related to this dataset, a larger study will be needed to validate the findings and to develop a convenient assay to apply in clinical practice. However, the significant infiltrate of IFN-gamma-producing cells sustains the biological plausibility of our results. Finally, the scRNA-seq did not include certain cell types such as neutrophils, likely because they did not survive the freeze-thaw process.

In summary, we demonstrated that the complex molecular biology in the kidney biopsy could be captured by the urine in patients with lupus nephritis. These processes provide insight into patient-specific disease pathogenesis that can address the issue of heterogeneity in SLE. These pathways can be tracked in the urine in real time and may not only improve diagnosis but also guide selection, dosing and monitoring of treatment, thus paving the way to develop a biologically grounded liquid biopsy.

## METHODS

### Patients and sample collection

This study enrolled SLE patient with urine protein to creatinine ratio > 0.5 undergoing clinically indicated renal biopsy, under protocols approved institutional review boards at each site and with informed consents. Only patients with a pathology report confirming lupus nephritis were included in the study. Renal biopsies were scored by a renal pathologist according to the International Society of Nephrology/Renal Pathology Society (ISN/RPS) guidelines and NIH activity and chronicity scales. Urine specimens were acquired at two clinical sites in the United States (Johns Hopkins University and New York University). The total urine volume was split into 2 50-ml Falcon tubes. Urine cells were pelleted by centrifugation at 200g for 10 min and the supernatant was aliquoted and stored at -80 °C.

### Urine Quantibody assay

The Kiloplex Quantibody protein array platform (QAH-CAA-X00, Raybiotech Life, Norcross, GA) was used for screening urine samples. The array was spotted with 1000 capture antibodies specific for 1000 different proteins in quadruplicate. All urine samples were clarified by centrifugation, and then diluted to yield a total protein concentration within the working range (500 ug/m - 1 mg/mL) before application to the arrays. Briefly, protein standards and urine samples were incubated on the array for 2 hours to allow the proteins to bind to the capture antibodies. A biotinylated antibody cocktail comprised of 1000 detection antibodies was subsequently added for incubation for 2 hours. Finally, streptavidin-Cy3 was added and left to incubate for 1 hour. Washing was performed between each step to remove the unbound reagents. After a final wash and dry, the slides were read with a fluorescent scanner, and data were extracted from the image using vendor-provided GAL file with a suitable microarray analysis software. Creatinine was measured for each urine sample (KGE005, R&D Systems, Inc., Minneapolis, MN), and all data were creatinine normalized before analysis. The complete list of the analytes measured in the urine is in the supplementary file “Kiloplex targets.csv”.

### Renal tissue single cell RNA sequencing

Renal tissue was collected, stored and processed as previously described (14). Briefly, research biopsy cores were collected from consented subjects as an additional biopsy pass or tissue from routine clinical passes. Only biopsies with confirmed lupus nephritis were included. Kidney tissue was frozen on site and shipped to a central processing location where it was thawed and disaggregated. Individual cells were retrieved and sorted by flow cytometry enriching for CD45+ leukocytes. For each single cell, the whole gene expression profile was sequenced using the CEL-Seq2 method (46). Data are publicly available (https://www.immport.org/resources/amp-ra-sle).

### Statistics

#### Principal component analysis

Urine protein concentrations were scaled (Z-normalized) within and across patients after log normalization. Scaling within samples favors the identification of the sample-specific pattern of expression regardless of the total urine protein concentration. Principal components were computed using the R Stats package version 3.5.2.

#### Pathway enrichment analysis

GSEA (47) was performed using the genes coding for the protein assessed by the Quantibody assay using the Gene Ontology Biological Process (48, 49). Pathways with at least 10 (but less than 500) genes represented in the Quantibody assay were included in the analysis. Enrichment p-values were computed based on 10,000 permutations and adjusted for multiple comparisons using the Benjamini-Hochberg procedure (i.e., false discovery rate) (50). The pathways captured by PC1 were determined based on the loading of each analyte on PC1.

## Data Availability

Data are publicly available.

https://www.immport.org/resources/amp-ra-sle

## AUTHOR CONTRIBUTION

A.F., M.P., J.B., and C.M. ideated the project. A.F, M.P, D.F, J.M., H.M.B., P.I., R.C, and B.D. acquired samples and supervised the processing. T.Z. and C.M. generated the proteomic data. A.F., M.P., J.B., D.F, J.M., H.M.B., P.I., R.C, A.A., D.A.R., C.C.B, N.H, D.W., and B.D contributed to the sample collection, processing, analysis and interpretation of the renal biopsy scRNAseq. A.F. devised the analytical plan, interpreted the results, and wrote the manuscript with the guidance of M.P, S.R., Y.Z., and L.M, and the contribution of J.B, D.A.R., R.C., H.M.B, and P.I. See Supplemental Acknowledgments for consortium details.

## ACKNOWLEDGMENTS

We thank the participating Accelerating Medicines Partnership network clinical sites and participants. We thank Ilya Korsunski, Maria Gutierrez, Fan Zhang, and Felipe Andrade for their critical review of the analytical plan and help with the interpretation of the results.

## SUPPLEMENTARY FILES

Kiloplex targets.csv

**Supplementary Figure 1.**
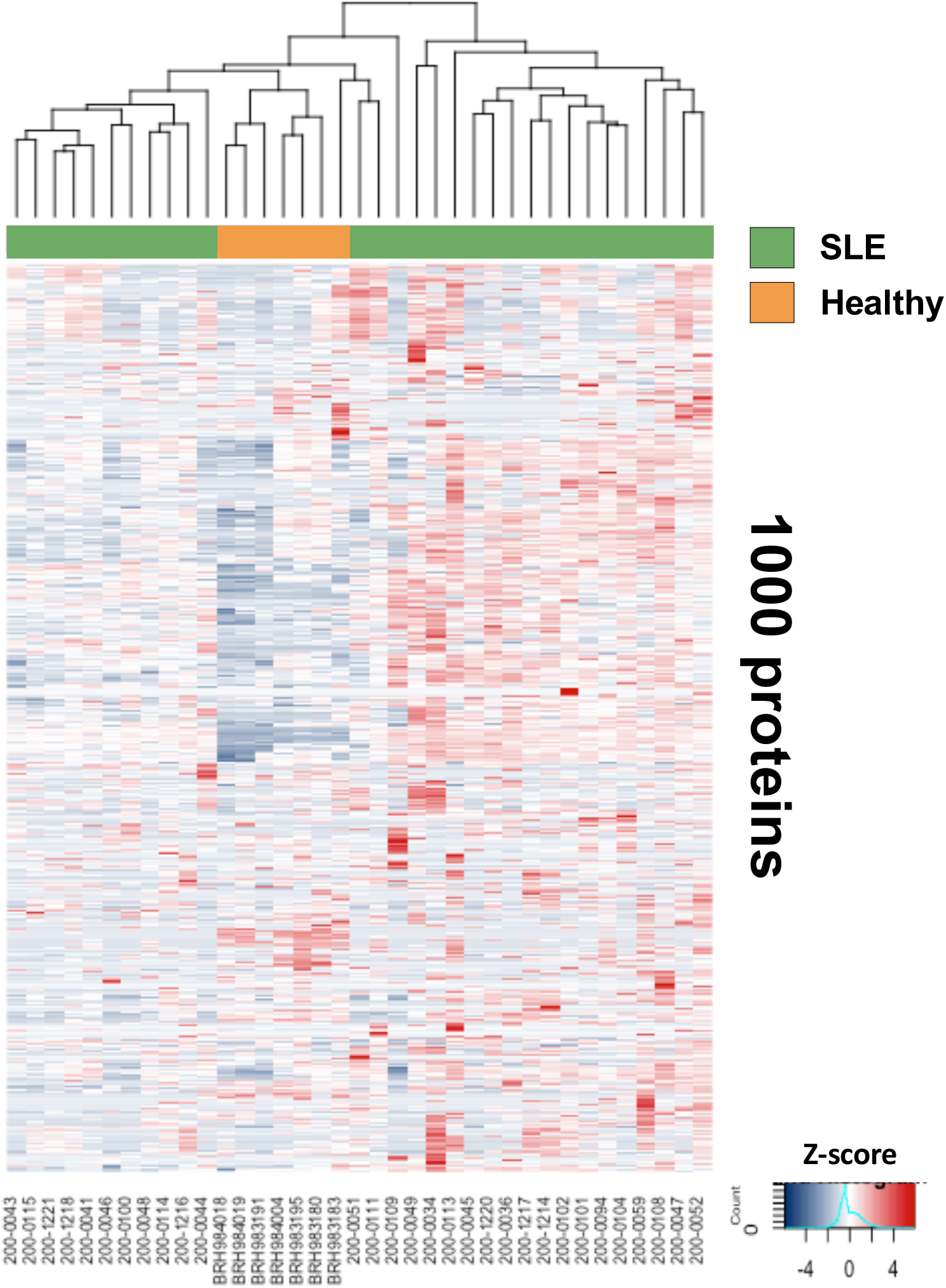
Overview of the urine biomarkers. Heatmap showing the relative concentration of the 1000 measure analytes (scaled by subject and analyte). Hierarchical clustering based on Ward’s rule and Euclidean distance.

**Supplementary Figure 2.**
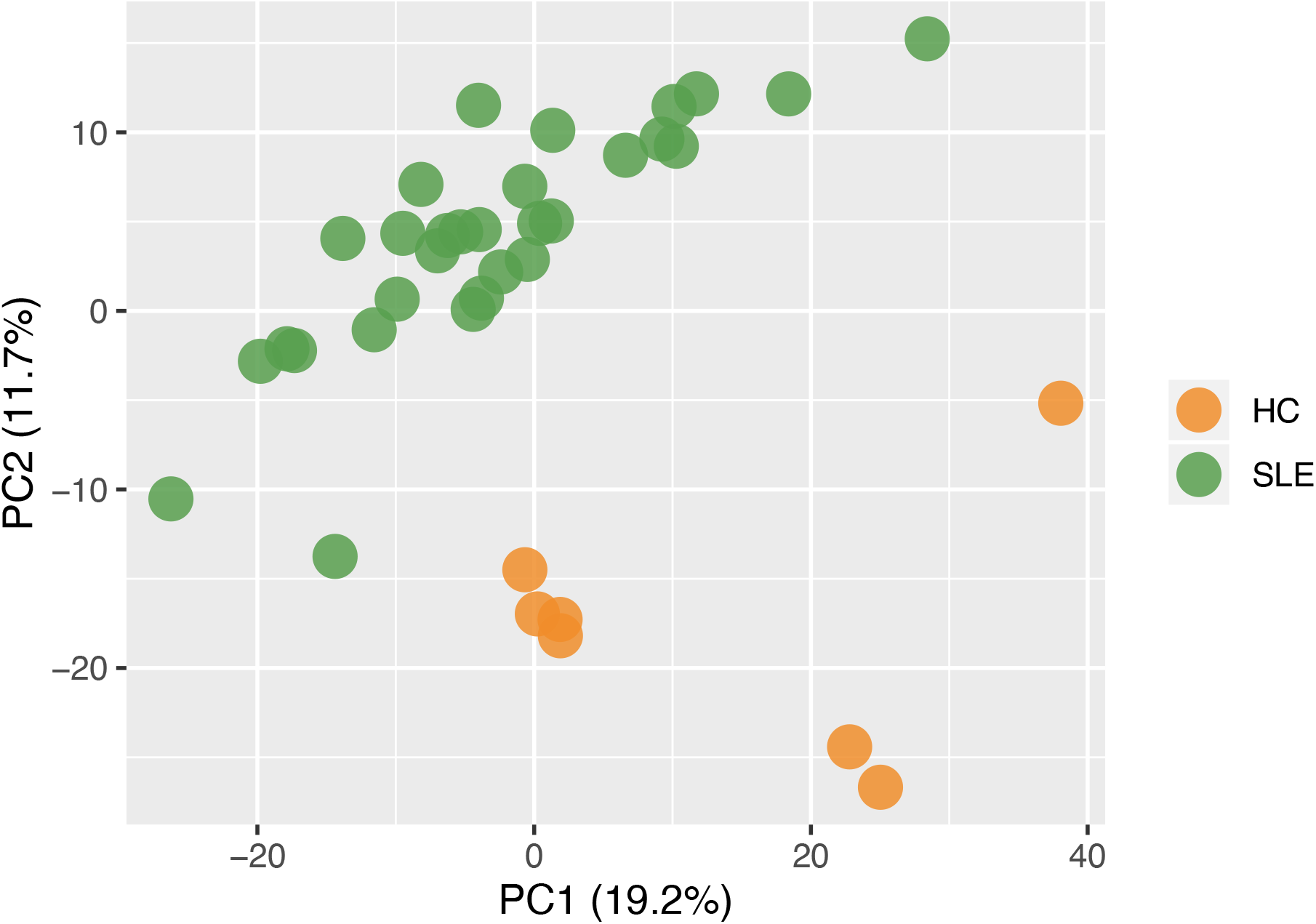
Principal component analysis of all urine analytes separates healthy subjects from lupus nephritis. PCA plot (n=37) of the first 2 principal components with % variance explained. Principal components are calculated independently in each analysis: PC1 in this figure does not correspond to PC in Figure 1.

**Supplementary Figure 3.**
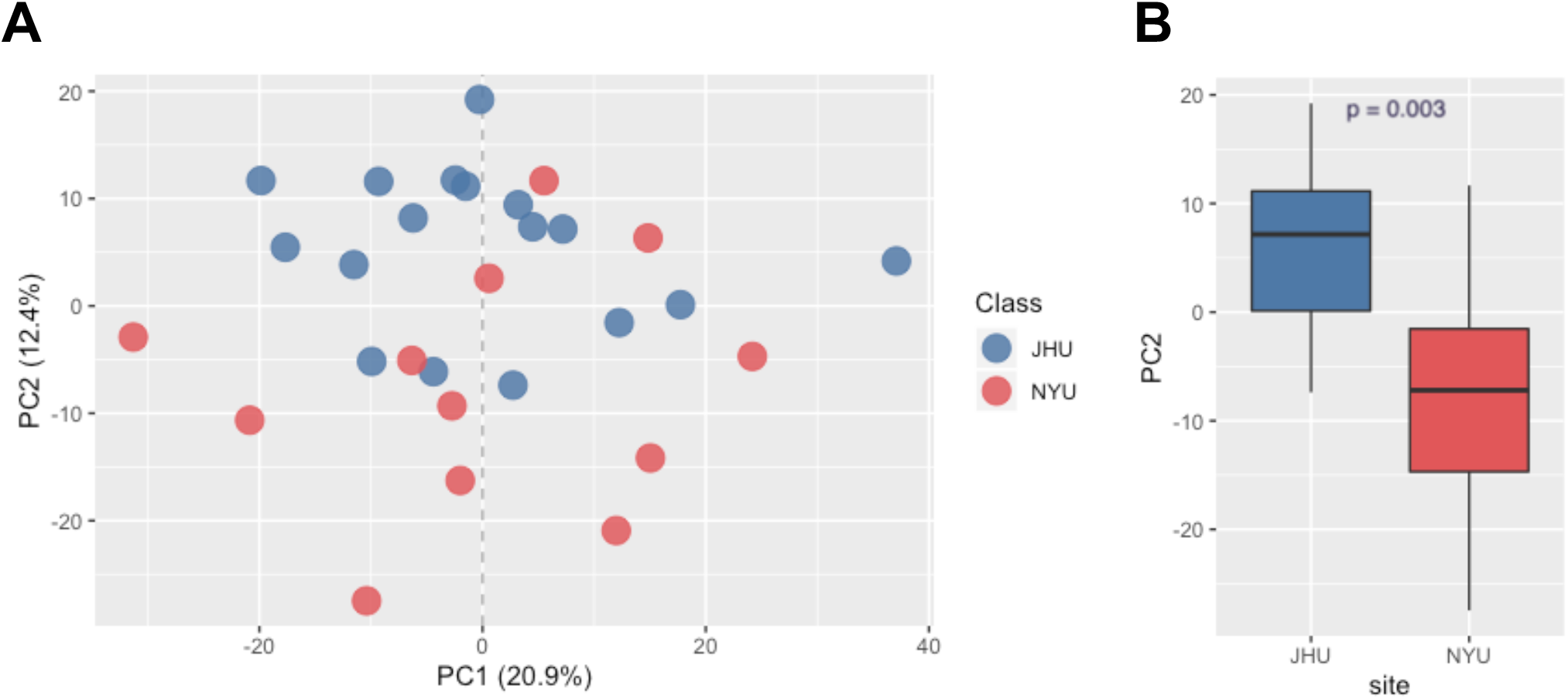
Principal component 2 detects collection site. (A-B) PCA (n=30) of the first 2 principal components of the urine proteome (% variance explained is indicated). PC2, but not PC1, was associated with the urine collection site. There was no biological pathway significantly enriched in PC2.

**Supplementary Figure 4.**
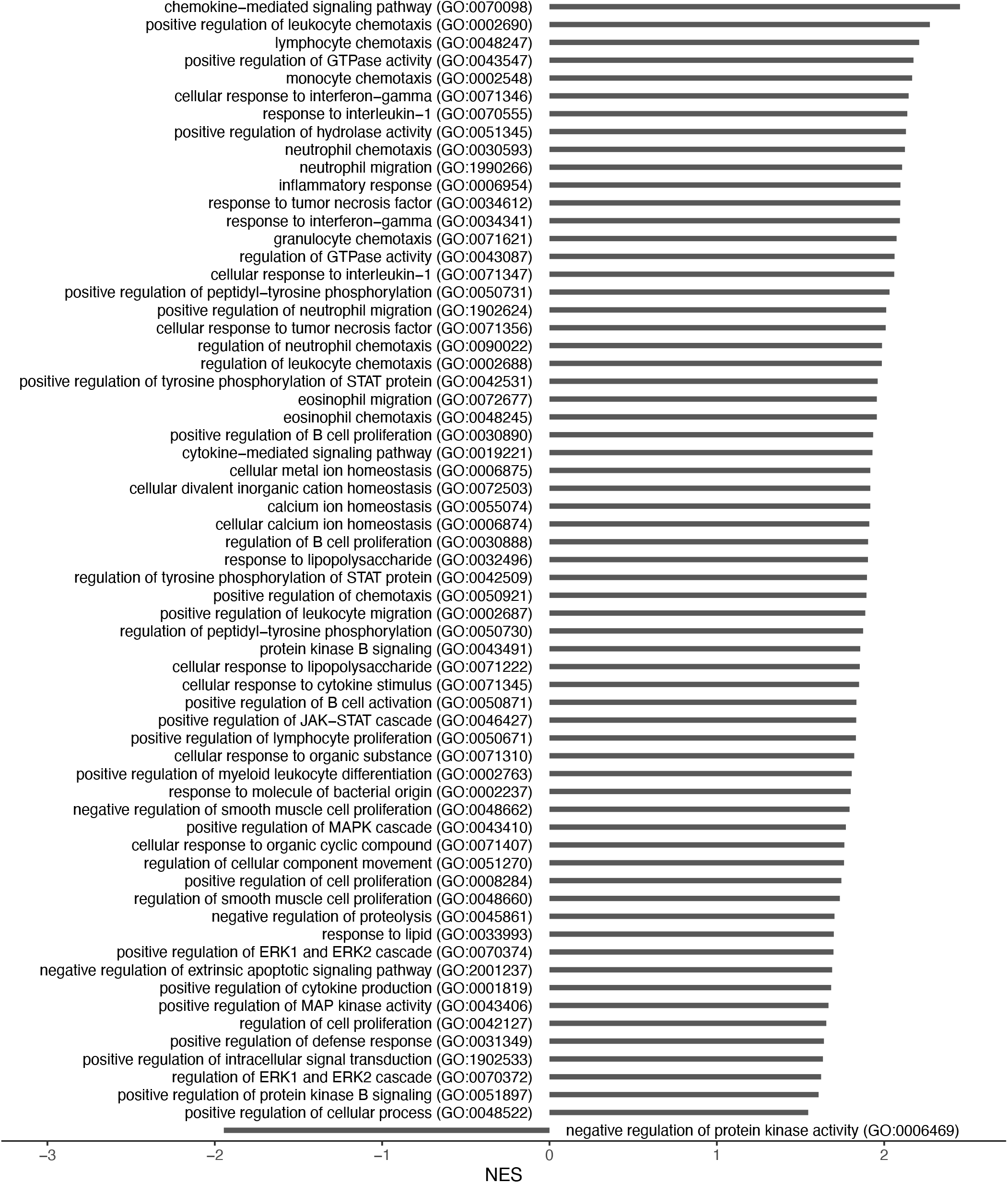
Complete list of pathways enriched in the urine proteome PC1. 2018 Gene Ontology Biological Process pathways with a FDR <5% are ordered by their normalized enrichment score.

**Supplementary Figure 5.**
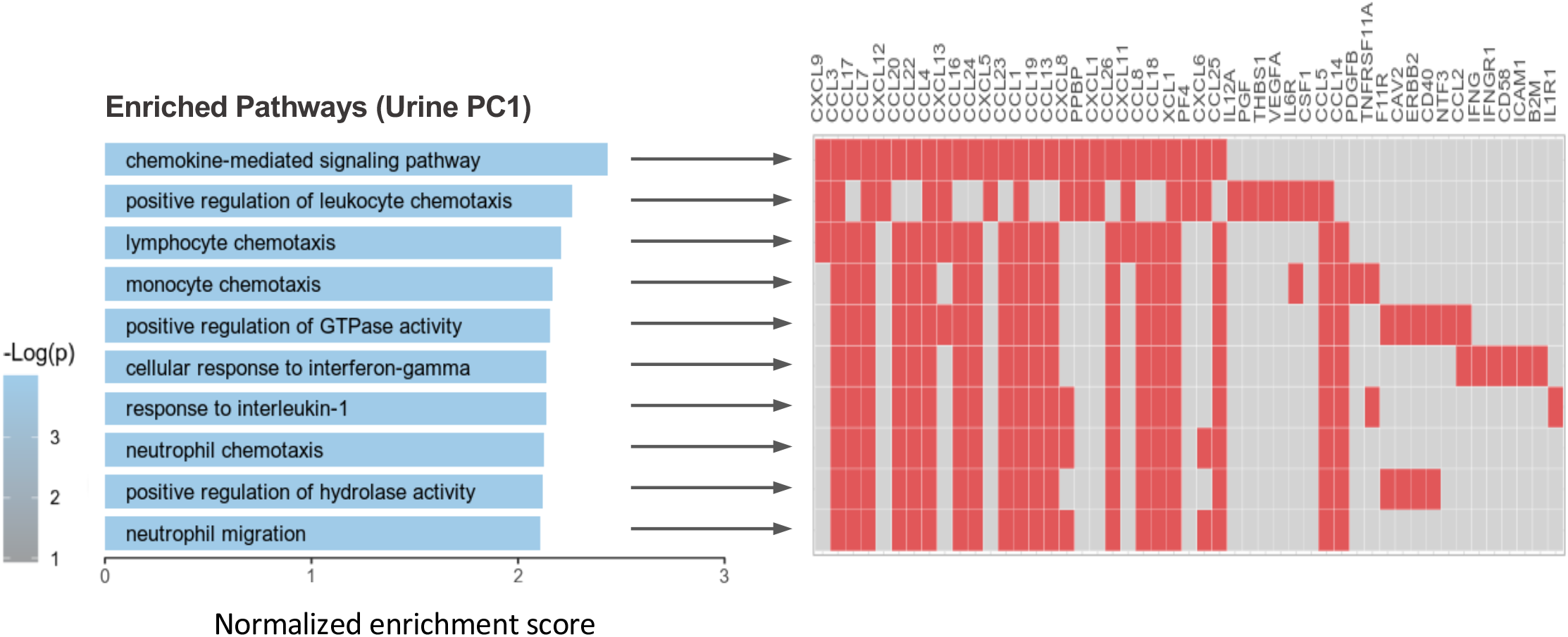
Definition of a chemokine score. First, the genes coding for the proteins defining the leading edge of “Chemokine-mediated signaling pathway GO:0070098”, the most enriched pathway in urine PC1 were identified. Then, a score for each cell was defined based on the sum of the normalized expression at the single cell level in the kidney. The same operation was repeated for “Cellular response to interferon-gamma (GO:0071346)”.

**Supplementary Figure 6.**
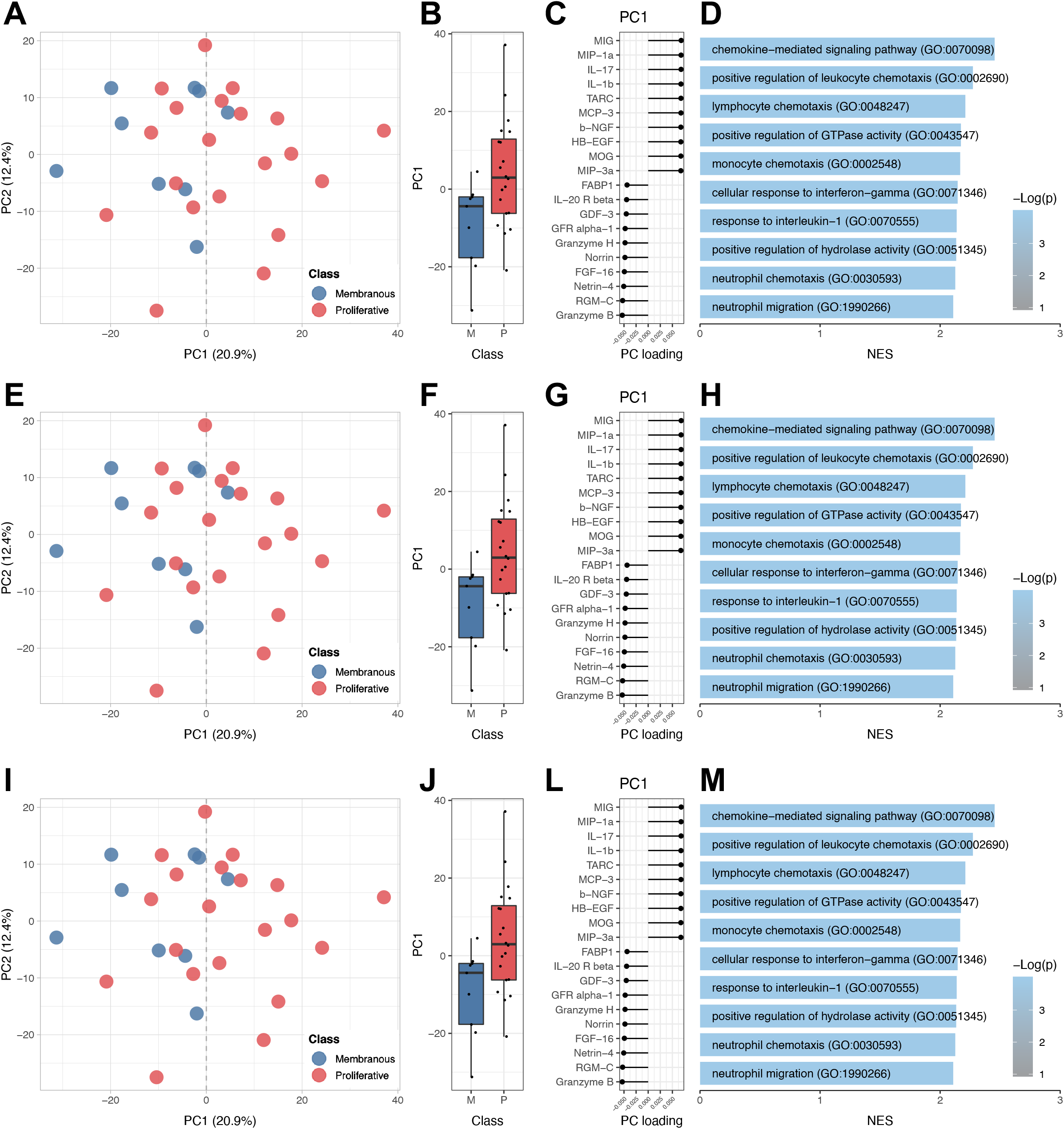
Adjustment for glomerular (albumin) or tubular (B2M) dysfunction. Urine analyte concentrations were normalized for the concentration of albumin (A-D), beta-2 microglobulin (E-H), or both (I-M) in order to account for non-specific protein excretion through the glomerulus, tubule, or both, respectively. Panels A,B, E, F, I, and J show the PCA (n=30) of the first 2 principal components of the urine proteome (% variance explained is indicated). Panels C, G, and L showed the top and bottom 10 PC1 loading values of the measured urine protein. Panels D, H, and M the top 10 enriched pathways PC1 using Gene Ontology Biological Process indicating the biological significance of PC1.

**Supplementary Figure 6.**
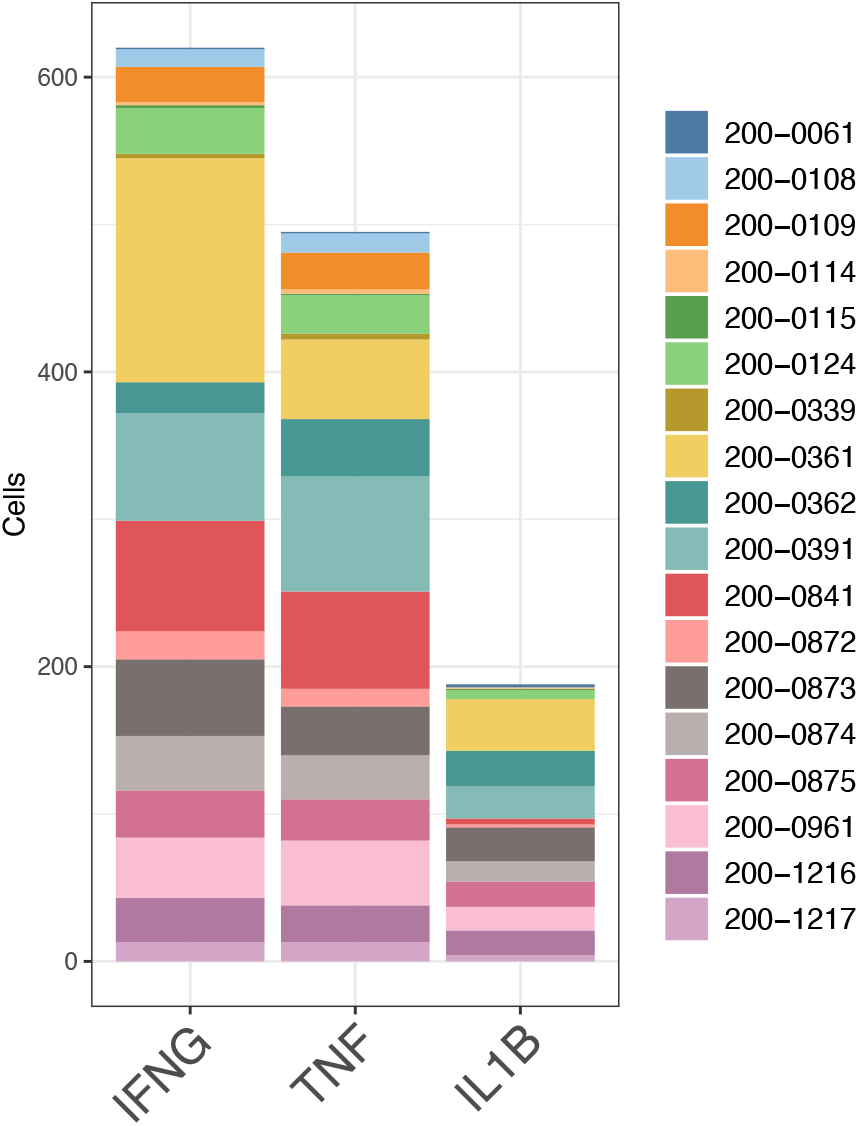
Distribution of cytokine positive cells among patients. Cells with positive cytokine UMIs were quantified in the renal biopsy single cell transcriptomics. The bar plot summarizes the individual contribution from samples with at least 10 high quality cells. As detailed in table S1, cytokine positive cells were found in all patients.

**Supplementary Figure 8.**
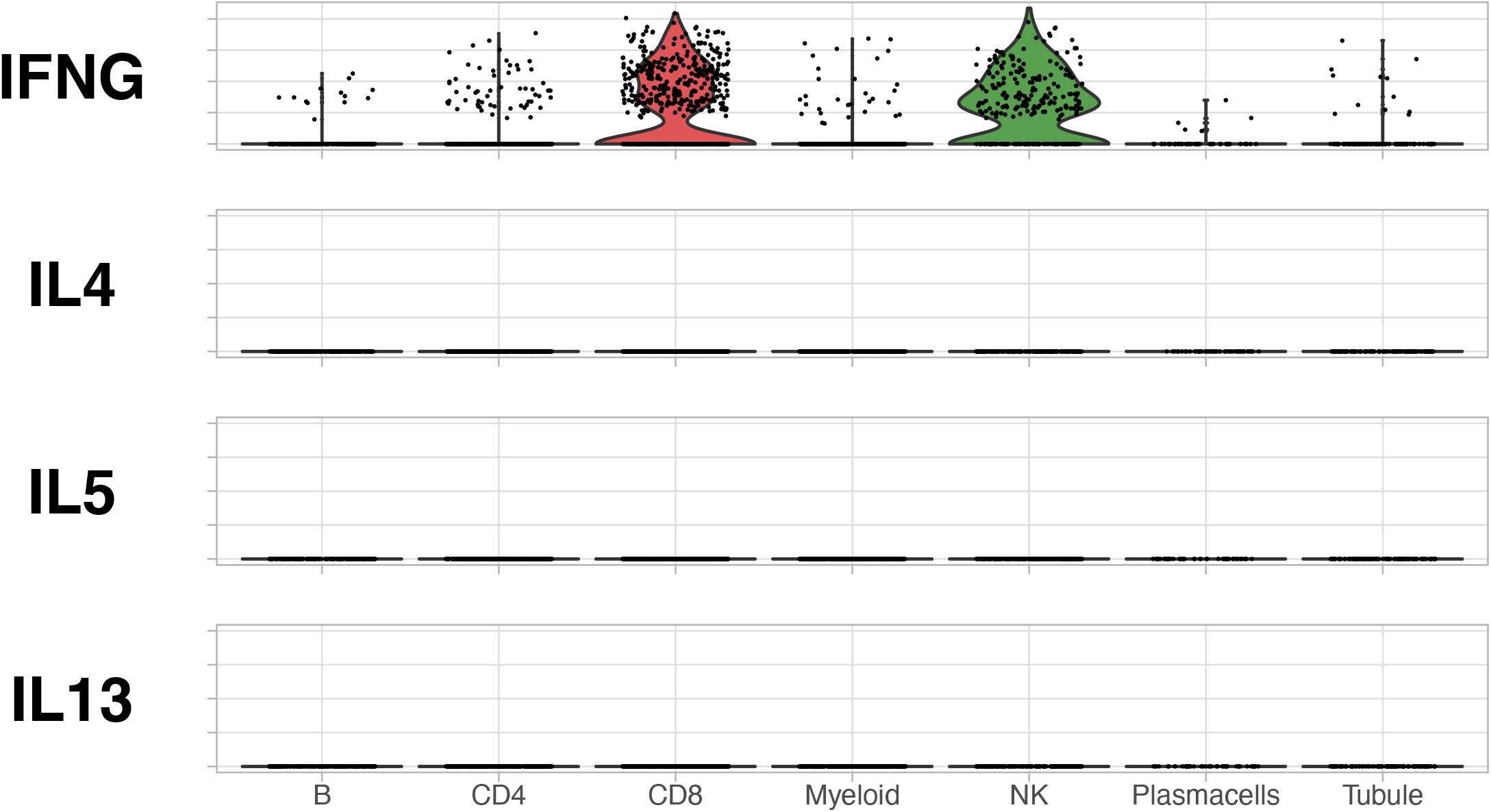
No expression of type 2 (Th2) immunity cytokines was detected in lupus nephritis kidney. Violin plots showing that there was abundant expression of the type 1 (Th1) immunity cytokine IFNG but none of the type 2 (Th2) immunity cytokines (IL4, IL5, and IL13) were detected in lupus nephritis kidney by single cell RNA sequencing.

